# Frequency of Atypical Pulmonary Manifestations of COVID-19 Patients on Chest CT-scan: A cross-sectional study

**DOI:** 10.1101/2023.07.28.23293261

**Authors:** Soheila Borji, Pouria Isavand, Mobin Azami, Ehsan Ghafouri

**Author notes:** **Corresponding author:** Ehsan Ghafouri, General practitioner, School of Medicine, Vali-e-Asr Hospital, Zanjan University of Medical Sciences, Zanjan, Iran, Postal code: 4515777978.

## Abstract

**Background:** Chest CT examination is significant in COVID-19 diagnosis due to its high sensitivity. Although typical chest CT findings have been discussed thoroughly in the literature throughout the pandemic, we aimed to investigate the prevalence of the atypical conclusions during the start of the Omicron variant insurgency and compare the results to studies conducted before its outbreak.

**Methods:** 606 confirmed COVID-19 cases were included in this study based on inclusion and exclusion criteria during January and February 2022. Demographic information of patients, including age and sex, was recorded. The computed tomography (CT) examination was carried out using a 100-slice scanner (Philips Brilliance 6 CT Scanner). One radiology attending and one resident evaluated SARS-CoV-2 RT-PCR-positive patients for atypical pulmonary CT findings. The obtained data were evaluated using R software version 4.1.1.

**Results:** 55% of patients were female, and the median age was 56 (IQR: 42, 69) 59% of patients had atypical findings on their pulmonary CT examination. These findings showed that pleural abnormalities were the most frequent atypical findings, with pleural thickening being the most common (17%). The double halo sign represented the least frequent atypical sign (0.2%).

**Conclusion:** Atypical findings were more prevalent in this study than its predecessors, while we acknowledge that other factors, such as study design and patient population, could have impacted it. The presence of atypical signs generally was not correlated with specific demographic groups, while some of these signs were more frequent in some groups.

## Introduction

In December 2019, the world encountered a new virus outbreak called coronavirus disease 2019 (COVID-19), leading to acute hypoxic respiratory failure and severe pneumonia [1-4]. According to the genomic sequencing analysis, this virus, identified as severe acute respiratory syndrome coronavirus 2 (SARS-CoV-2), belongs to the Orthocoronavirinae subfamily of Coronaviridae and mainly affects the upper respiratory tract [5, 6]. Several viruses in this subfamily, including the Middle East Respiratory Disease coronavirus and Severe Acute Respiratory Syndrome coronavirus, are well known for being the primary causes of highly contagious diseases (MERS-CoV)[5, 7, 8].

Since the pandemic’s start, SARS-CoV-2 has evolved into new variants of concern, according to its genomic sequencing analysis, with different virulence, infectiousness, and clinical picture. On 26 November 2021, WHO designated B.1.1.529 as a variant of concern and named it “Omicron.” As of 6 January 2022, the variant has been confirmed in 149 countries. Following the original BA.1 variant, several subvariants of Omicron have emerged: BA.2, BA.3, BA.4, and BA.5. Omicron (BA.1) has more mutations than any previous SARS-CoV-2 variant with 50 mutations[9].

The most frequent symptoms of COVID-19 are fever, dry cough, dyspnea, sore throat, and fatigue. Besides, other symptoms have been observed, including abdominal pain, vomiting, and diarrhea [8, 10]. Multiple studies have reported nonspecific or atypical symptoms in some COVID-19 patients [11].

Besides the clinical manifestation described above, the gold-standard method for diagnosing the SARS-CoV-2 infection is reverse transcription polymerase chain reaction (RT-PCR). However, the physician may use other paraclinical and diagnostic measures for early detection and to identify the extent of lung involvement. [12-14]. Pulmonary computed tomography has been reported to have 97% sensitivity [15]. Thus, the obtained data in pulmonary CT examination plays an essential role in disease monitoring and the subsequent inquiry into the effectiveness and adequacy of our therapeutic approach [16]. According to several studies, the most common reported CT lesions to include reversed halo sign, vascular enlargement, crazy paving patterns, reticular patterns, air-bronchogram, airway changes, and pleural changes. [12].

Furthermore, vascular dilation, air bubble sign, subpleural lines, halo and reverse halo sign, and bronchial dilatation have been reported to be accepted as the radiologic signs seen in COVID-19 patients [17]. Unlike the typical signs of COVID-19 on chest CT scans, which have been well demonstrated in different studies, the atypical manifestations have not been paid attention to the same extent in the “Omicron era.” In a review article, researchers have summarized the atypical chest CT findings of COVID-19 patients as, Central involvement, Isolated upper lobe involvement, Solitary involvement, Peribronchovascular involvement, Lobar consolidation, Tree-in-bud pattern, Nodules, Pleural effusion, Pericardial effusion, Subpleural sparing, and White lung [17]. Therefore, this study aimed to investigate the atypical pulmonary manifestations in COVID-19 patients and their frequency in the patient population during the first two months of 2022, after WHO designated Omicron as a variant of concern.

### Study design and data collection

A cross-sectional study was performed on COVID-19 patients during January and February 2022 at Vali-e-Asr and Mousavi Hospitals in Zanjan, Iran. The RT-PCR-positive patients were re-evaluated to meet the inclusion and exclusion criteria: having no lung disease and history of lobectomy, patients older than 18, and not pregnant women. After applying the mentioned criteria, 606 patients were selected for investigation through chest CT examination, irrespective of disease severity and stage. The Iran National Committee for Ethics in Biomedical Research approved this study with the ethics code of IR.ZUMS.REC.1401.175. Subsequently, CT images were obtained for each patient.

### CT imaging protocol

The CT examination was carried out using a 100-slice scanner (Philips Brilliance 6 CT Scanner) in a supine position with a single breath-hold. The CT imaging protocol parameters were as follows: scan direction (craniocaudally), tube voltage (120kV), tube current (100 mA), slice collimation (6 × 1.5 mm), pitch (1.5), rotation time (0.75 s), and scan time (18.02 s). Finally, slice thickness and an interval were set as 3 mm to reconstruct images.

### CT imaging interpretation

Obtained CT images were evaluated independently by a senior attending with experience of 7 years in practice and a third-year radiology resident. Initially, all patients were assessed for the presence or absence of atypical signs. In this study, (1) isolated upper lobe involvement, (2) solitary involvement, (3) peribronchovascular involvement, (4) lobar consolidation, (5) tree-in-bud pattern, (6) centrilobular nodules, (7) pleural effusion, (8) pleural thickening, (9) pericardial effusion, (10) subpleural sparing, (11) white lung, (12) halo sign, (13) reverse halo sign, (14) double halo sign, (16) target shaped opacity, (17) lymphadenopathy, (18) cavitation, (19) air bubble sign, and (20) airway changes, were evaluated as atypical findings.

### Statistical analysis

The R software 4.1.1 was used to analyze the data. Clinical findings, patient age, and sex were all recorded in terms of relative frequency and percentage. The age is considered to be median (IQR). Pearson’s Chi-squared test and Fisher’s exact test were used to compare atypical results by sex. Patients’ ages were also classified into three age groups: less than 40, 40 to 60, and more than 60. They also were compared regarding atypical findings using Fisher’s exact test. Some of the cases are shown in the figures.

## Results

### Demographics and atypical findings

The demographics and atypical findings of evaluated patients in our study are shown in Table 1. According to obtained results, 55% of patients were female, and the median age was 56 (IQR: 42, 69) years. Patients more than 60 years of age (42%) were shown to have some atypical findings on their pulmonary CT examination more frequently. It has been demonstrated that 59% of all investigated patients had atypical manifestations. The pleural thickening was the most frequent atypical finding (17%) in COVID-19 patients, while the double halo sign represented the least frequent atypical sign (0.2%). No patient in our study exhibited any evidence of cavitation on their lung CT scan.

**Table 1.** Demographics and the frequency of atypical findings.

| Variable | N (total=606)<br><sub>1</sub> |
| --- | --- |
| Sex |  |
| Female | 332 (55%) |
| Male | 274 (45%) |
| Age | 56 (42, 69)<br><sub>2</sub> |
| Atypical Findings | 355 (59%) |
| Isolated Upper Lobe Involvement | 10 (1.7%) |
| Solitary Involvement | 17 (2.8%) |
| Peribronchovascular Involvement | 43 (7.1%) |
| Lobar Consolidation | 3 (0.5%) |
| Tree in bud Pattern | 3 (0.5%) |
| Centrilobular Nodules | 13 (2.1%) |
| Pleural Effusion | 39 (6.4%) |
| Pleural Thickening | 103 (17%) |
| Pericardial Effusion | 18 (3.0%) |
| Subpleural Sparing | 29 (4.8%) |
| White Lung | 15 (2.5%) |
| Halo sign | 42 (6.9%) |
| Reverse Halo Sign | 59 (9.7%) |
| Double Halo Sign | 1 (0.2%) |
| Target Shaped Opacity | 28 (4.6%) |
| Lymphadenopathy | 29 (4.8%) |
| Cavitation | 0 (0%) |
| Air Bubble Sign | 14 (2.3%) |
| Airway Changes | 102 (17%) |
| Age group |  |
| (0,40yrs] | 145 (24%) |
| (40,60yrs] | 208 (34%) |
| (more than 60yrs) | 253 (42%) |

<sub>1</sub> N: number of patients (%)
<sub>2</sub> Median (IQR= Interquartile range )

### Atypical findings according to sex and age

In this part, we investigated the prevalence of atypical findings according to sex and age. The presence of atypical findings on lung CT generally did not significantly differ in men compared to women (Table 2). Moreover, our analysis demonstrated that peribronchovascular involvement (p=0.003), pericardial effusion (p=0.013), and reverse halo sign (p=0.008) were significantly more frequent in women than men. Meanwhile, pleural thickening (p=0.023) and subpleural sparing (p=0.008) were more common in men (Table 2).

**Table 2.** Atypical findings according to sex.

| CT findings | Female, N = 332<sub>1</sub> | Male, N = 274<sub>1</sub> | p-value<sub>2</sub> |
| --- | --- | --- | --- |
| Atypical Findings | 192 (58%) | 163 (59%) | 0.7 |
| Isolated Upper Lobe Involvement | 5 (1.5%) | 5 (1.8%) | 0.8 |
| Solitary Involvement | 9 (2.7%) | 8 (2.9%) | 0.9 |
| Peribronchovascular Involvement | 33 (9.9%) | 10 (3.6%) | 0.003 |
| Lobar Consolidation | 2 (0.6%) | 1 (0.4%) | >0.9 |
| Tree in bud Pattern | 0 (0%) | 3 (1.1%) | 0.092 |
| Centrilobular Nodules | 5 (1.5%) | 8 (2.9%) | 0.2 |
| Pleural Effusion | 19 (5.7%) | 20 (7.3%) | 0.4 |
| Pleural Thickening | 46 (14%) | 57 (21%) | 0.023 |
| Pericardial Effusion | 15 (4.5%) | 3 (1.1%) | 0.013 |
| Subpleural Sparing | 9 (2.7%) | 20 (7.3%) | 0.008 |
| White Lung | 7 (2.1%) | 8 (2.9%) | 0.5 |
| Halo sign | 20 (6.0%) | 22 (8.0%) | 0.3 |
| Reverse Halo Sign | 42 (13%) | 17 (6.2%) | 0.008 |
| Double Halo Sign | 1 (0.3%) | 0 (0%) | >0.9 |
| Target Shaped Opacity | 11 (3.3%) | 17 (6.2%) | 0.092 |
| Lymphadenopathy | 16 (4.8%) | 13 (4.7%) | >0.9 |
| Cavitation | 0 (0%) | 0 (0%) |  |
| Air Bubble Sign | 7 (2.1%) | 7 (2.6%) | 0.7 |
| Airway Changes | 52 (16%) | 50 (18%) | 0.4 |

<sub>1</sub> N: number of patients (%)
<sub>2</sub> Pearson's Chi-squared test; Fisher's exact test

The frequency of atypical manifestations based on age categories was studied. Patients below 40 years of age, 40-60 years, and more than 60 represented 61%, 55%, and 60% positive for atypical signs, respectively, with no statistically significant correlation between the existence of atypical findings in general and patients’ age (p=0.5). Detailed analysis indicated that solitary involvement (p=0.018), halo sign (p=0.004), reverse halo sign (p<0.001), target-shaped opacity (p=0.011), pleural effusion (p<0.001), pleural thickening (p<0.001), white lung (p<0.001), lymphadenopathy (p<0.001), airway changes (p<0.001) and pericardial effusion (p=0.040) were significantly different between age categories with the later six being more frequent in patients above 60 years of age (Table 3).

**Table 3.** Atypical findings in regards to patient’s age.

| CT findings | (0,40yrs], N = 145<sub>1</sub> | (40,60yrs], N = 208<sub>1</sub> | >60yrs, N = 253<sub>1</sub> | p-value<sub>2</sub> |
| --- | --- | --- | --- | --- |
| Atypical Findings | 89 (61%) | 115 (55%) | 151 (60%) | 0.5 |
| Isolated Upper Lobe Involvement | 3 (2.1%) | 4 (1.9%) | 3 (1.2%) | 0.8 |
| Solitary Involvement | 9 (6.2%) | 5 (2.4%) | 3 (1.2%) | 0.018 |
| Peribronchovascular Involvement | 11 (7.6%) | 10 (4.8%) | 22 (8.7%) | 0.3 |
| Lobar Consolidation | 1 (0.7%) | 2 (1.0%) | 0 (0%) | 0.3 |
| Tree in bud Pattern | 1 (0.7%) | 1 (0.5%) | 1 (0.4%) | >0.9 |
| Centrilobular Nodules | 3 (2.1%) | 3 (1.4%) | 7 (2.8%) | 0.6 |
| Pleural Effusion | 6 (4.1%) | 4 (1.9%) | 29 (11%) | <0.001 |
| Pleural Thickening | 12 (8.3%) | 26 (12%) | 65 (26%) | <0.001 |
| Pericardial Effusion | 2 (1.4%) | 3 (1.4%) | 13 (5.1%) | 0.040 |
| Subpleural Sparing | 8 (5.5%) | 14 (6.7%) | 7 (2.8%) | 0.12 |
| White Lung | 2 (1.4%) | 0 (0%) | 13 (5.1%) | <0.001 |
| Halo sign | 17 (12%) | 17 (8.2%) | 8 (3.2%) | 0.004 |
| Reverse Halo Sign | 19 (13%) | 29 (14%) | 11 (4.3%) | <0.001 |
| Double Halo Sign | 0 (0%) | 1 (0.5%) | 0 (0%) | 0.6 |
| Target Shaped Opacity | 10 (6.9%) | 14 (6.7%) | 4 (1.6%) | 0.011 |
| Lymphadenopathy | 3 (2.1%) | 4 (1.9%) | 22 (8.7%) | <0.001 |
| Cavitation | 0 (0%) | 0 (0%) | 0 (0%) | - |
| Air Bubble Sign | 5 (3.4%) | 6 (2.9%) | 3 (1.2%) | 0.2 |
| Airway Changes | 10 (6.9%) | 26 (12%) | 66 (26%) | <0.001 |

<sub>1</sub> N: number of patients (%)
<sub>2</sub> Pearson's Chi-squared test; Fisher's exact test

## Discussion

Due to the high sensitivity of pulmonary computed tomography examination in diagnosing SARS-CoV-2 infections, it is essential to investigate its disparate radiologic manifestations. Contrary to the typical findings which have been robustly canvassed since the start of the pandemic, atypical manifestations generate a significant problem in diagnosis, and their prevalence can potentially vary with each variant of concern. In this study, we evaluated atypical findings in pulmonary CT examination in confirmed COVID-19 patients in the first two months of 2022 after WHO designated Omicron as a variant of concern. The results showed that 59% of patients had at least one atypical sign on their lung CT. On the other hand, it was revealed that atypical findings, in general, were not significantly different based on sex and age. However, it was shown that the type of atypical radiologic signs could vary in patients regarding age and sex.

Compared to previous studies, our findings represented more incidence of atypical pulmonary signs in COVID-19 patients [18, 19]. Gurumurthy et al. [19] have investigated typical pulmonary manifestations of COVID-19 in India, reported the prevalence of atypical signs as 21.1% out of 298 cases, and reported that pulmonary cysts (9%) were the most frequent pulmonary atypical manifestation. Here, we demonstrated that pleural thickening was the most frequent atypical finding (17%), with 59% of SARS-CoV2 RT-PCR positive patients having at least one atypical radiologic sign on their lung CT imaging. Moreover, Haghighi-Morad et al. [18] have investigated the Atypical presentation of COVID-19 and have stated that 3% of patients had atypical findings on their lung CT. In both of these studies, contrary to our findingss, the frequency of atypical pulmonary CT manifestations in male patients was higher than in female patients. These discrepant findings may be caused by variations in sample size, patient characteristics, epidemiologic variables in specific populations, disease severity, the number and variety of atypical parameters evaluated across studies, and most importantly, the variant that is identified as the primary variant of concern during the study. In a systematic review and meta-analysis conducted by Zarifian et al. [20] pleural thickening (33.35%) and bronchial wall thickening (15.48%), were major atypical lung CT findings, with other signs such as pleural effusion (6.96%), lymphadenopathy (5.19%), cavitation (1.1%), and pneumothorax (0.89%) being other atypical radiologic manifestations investigated, all similar to our findings. The major discrepancy, was Halo sign which was not deemed as an atypical finding and its prevalence was much higher (25.63%).

In another study, Korkmaz et al. [21] have shown that the most common atypical finding in COVID-19 patients was nodular lesions. However, they have revealed that the atypical results can vary due to the pandemic period as they showed that central distribution, as an atypical manifestation, was significantly different between those infected during the first six months of the pandemic (March to August 2020) and in the second (September 2020 to February 2021) (p=0.001). This information can be summarized by asserting that the frequency of atypical radiologic signs in COVID-19 patients relies on a number of variables, including but not limited to the variation of concern.

Awareness of CT findings, including atypical manifestations, can be considered an effective implementation to prevent misdiagnoses of COVID-19 patients. We showed that 59% of investigated patients were positive for atypical findings. The pleural abnormalities were shown to be the most frequent atypical manifestation (23.4%) in RT-PCR-positive patients in our study. As Saha et al. [22] discussed, various types of pleural involvement and their frequencies vary depending on disease severity and stage, as pleural thickening and pleural retraction are more common and have been more commonly attributed to the early disease. In contrast, the prevalence of pleural effusion is much lower, and pneumothorax is uncommon. Our results are similar to previous studies regarding the frequency of pleural involvement. Bai et al. [23] have investigated the frequency of pleural lesions in 219 patients with COVID-19 and reported that common pleural abnormality among COVID-19 patients was pleural thickening (15%) followed by pleural effusion (4%), which is congruent with our findings. Recent studies have indicated the importance of pleural abnormalities in the disease outcome. Hence, it has been shown that patients with pleural effusion have more pulmonary inflammatory responses due to refractory disease [24]. More interestingly, Hussein et al. [25] have suggested that effusion development may have been solely related to the virus.

## Conclusion

This study investigated the frequency of atypical pulmonary CT manifestations in COVID-19 patients during January and February of 2022 after WHO designated Omicron as a variant of concern. Our results showed that the prevalence of atypical pulmonary manifestations in COVID-19 patients was 59% in general. The pleural abnormalities were the most frequent atypical findings in the studied patients. Given the importance of these manifestations in misdiagnosis and quarantine delays, they should be given more attention in pulmonary CT scans. They could be more prevalent in new variants of concern.

## Data Availability

All data produced in the present study are available upon reasonable request to the authors

## Abbreviations

COVID-19: Coronavirus disease 2019
SARS-CoV-2: Severe acute respiratory syndrome coronavirus 2
SARS-CoV-1: Severe Acute Respiratory Syndrome coronavirus-1
RT-PCR: Reverse transcription polymerase chain reaction
CT-scan: Computed tomography scan

---

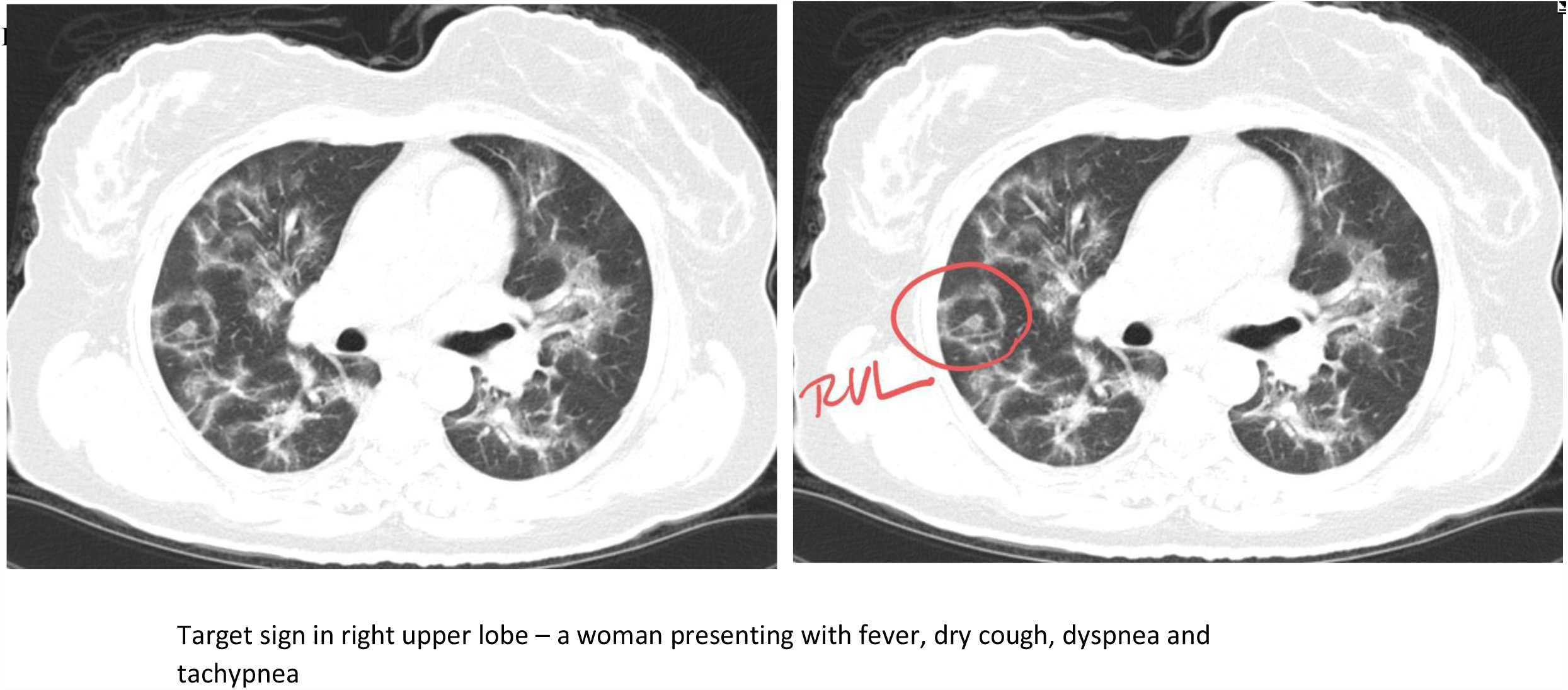

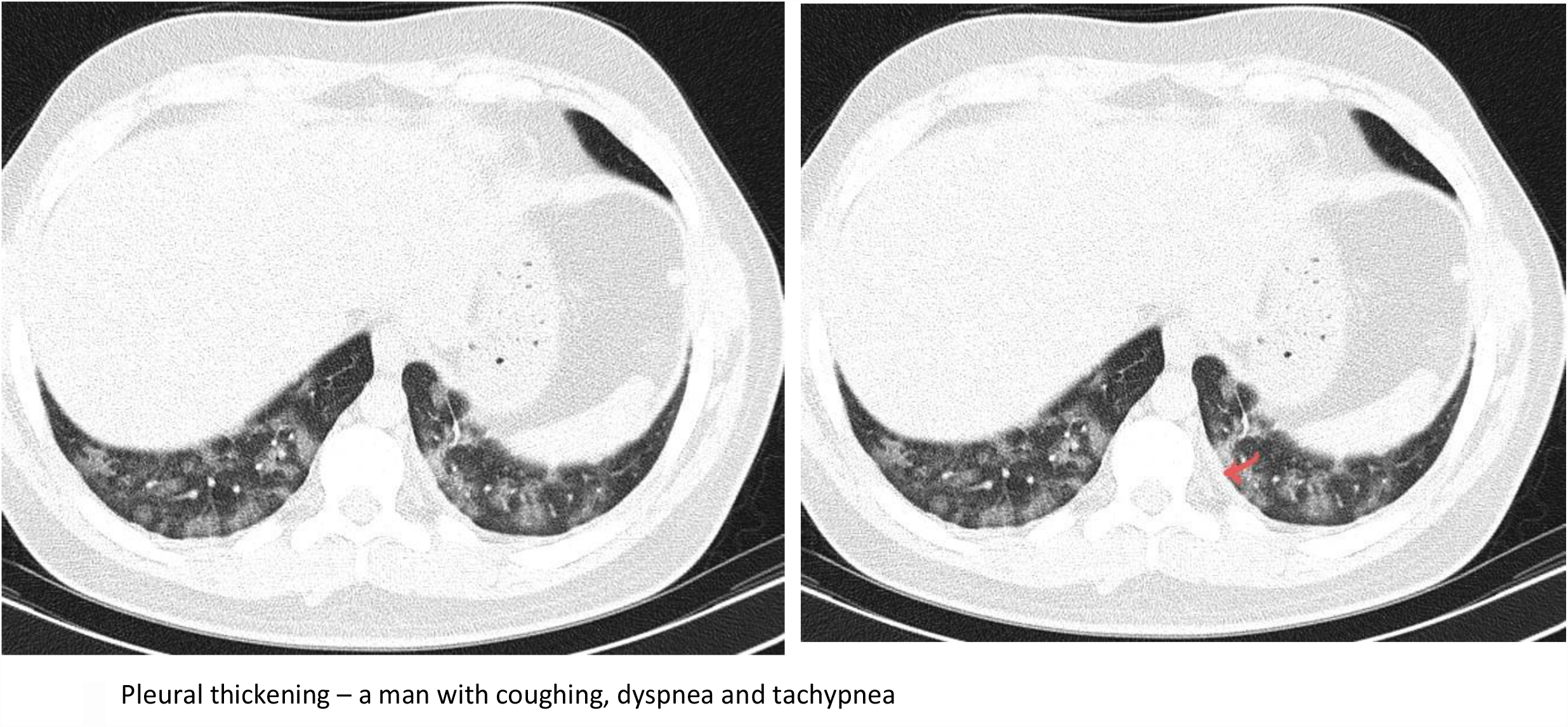

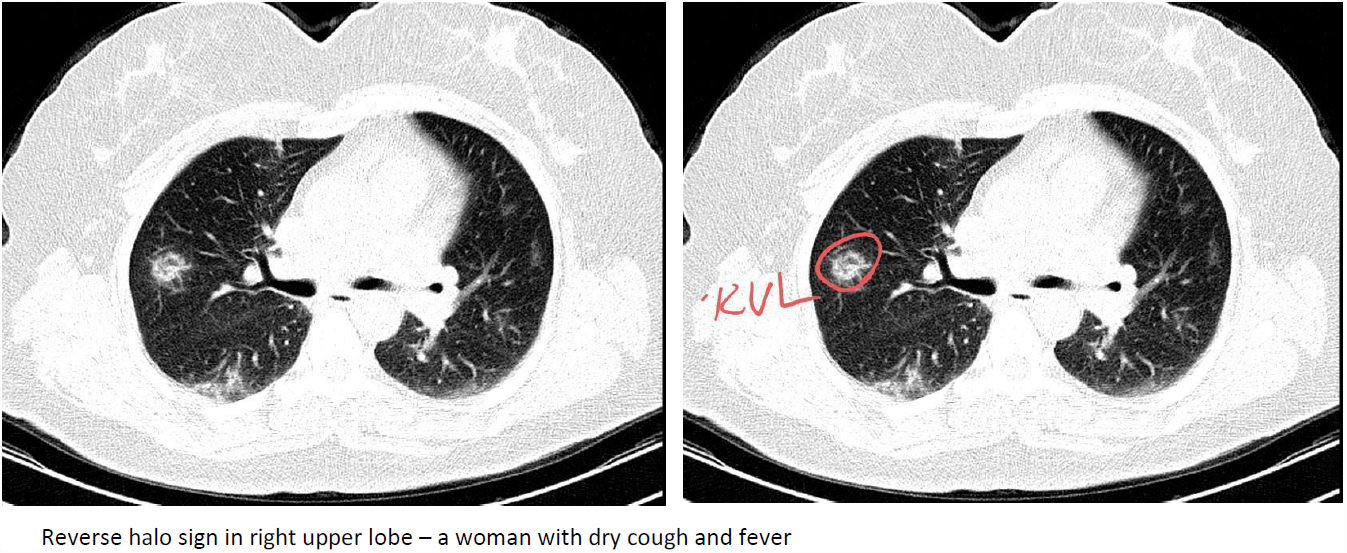

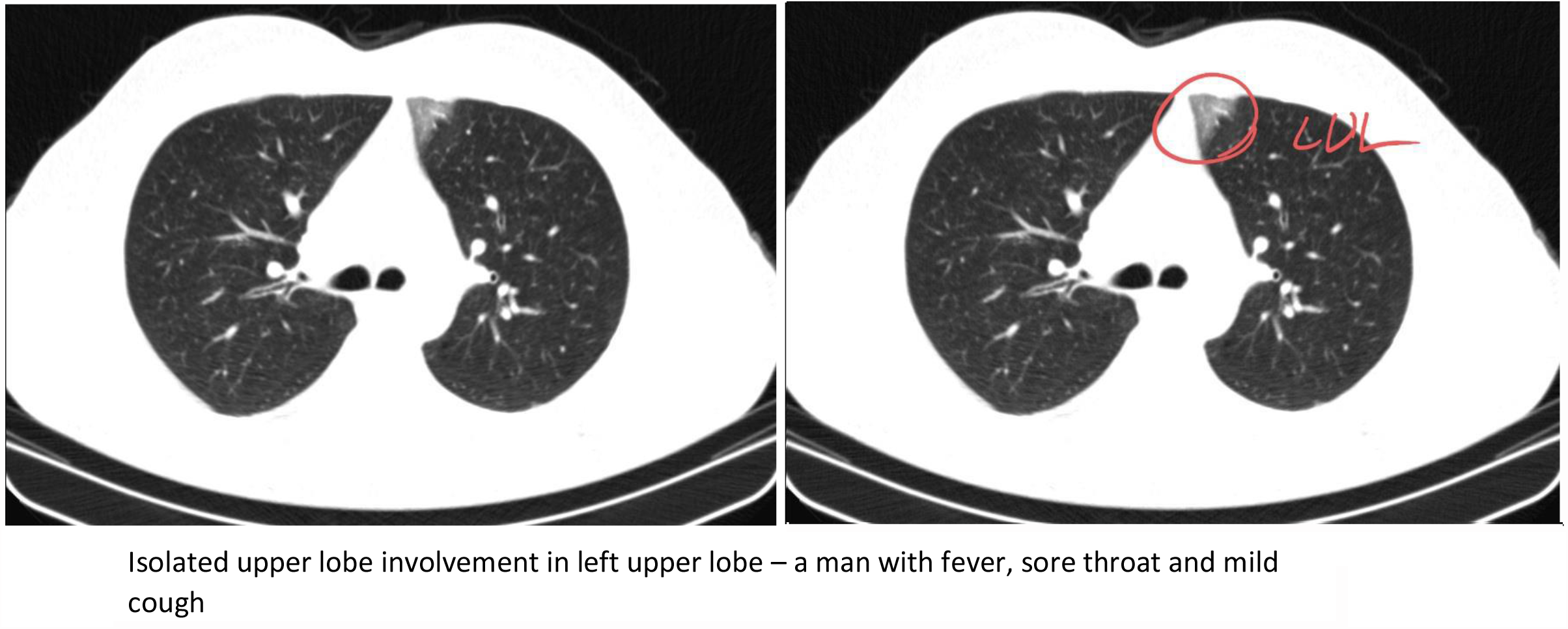

